# Background styles in systematic review articles: a cross-sectional study protocol

**DOI:** 10.1101/2020.01.19.20018127

**Authors:** Yuki Kataoka, Shunsuke Taito, Sachiko Yamamoto-Kataoka, Yasushi Tsujimoto, Hajime Yamazaki, Toshi A. Furukawa

## Abstract

**Background:** The background section of a medical journal article has the important function to communicate readers the value of the research question. However, little is known about how authors describe their “niche” to emphasize the importance of their research question. This study aims to examine the methods the authors use in order to delineate their niche in systematic reviews (SR).

**Methods:** We will conduct a cross-sectional study. We will include Original SR articles published in top 50 journals in MEDICINE, GENERAL & INTERNAL category in Journal Citation Reports 2018. We will conduct content analysis of background sections. The primary outcome will be whether the article was published in top 10 journal or not. We will use chi-squared test and logistic regression analysis. The primary analysis will be logistic regression predicting publication in high impact journals, with covariates. Two-tailed p values will be considered statistically significant if less than 0.05. Discussion: This is the first study to investigate the influence of what to present and not present in the backgrounds section to be accepted in the highly cited journals among SR articles.

## Backgrounds

The background section of a medical journal article has the important function to communicate readers the value of the research question. There are many textbooks and review articles on how to write it based on expert opinions (1–3). In addition, there are several analyses that examined its structure in medical research articles. The basic structure of the background section may be characterized as follows: “establishing a territory”, “establishing a niche”, and “occupying the niche” (4,5).

However, little is known about how authors describe their “niche” to emphasize the importance of their research question. This study aims to examine the methods the authors use in order to delineate their niche in systematic reviews (SR). We have focused on SRs because SRs are the most important research design, in terms of practicing evidence-based medicine (6).

## Meshods

### Protocol

We followed the reporting guideline of meta-epidemiological study to prepare this protocol (7). We will publish this protocol in medRxiv (https://www.medrxiv.org/).

### Study design

We will conduct a cross-sectional study.

### Eligibility criteria

Original SR articles published in top 50 journals in MEDICINE, GENERAL & INTERNAL category in Journal Citation Reports 2018 (8). We show the lists in table 1. We will include articles published in 2018. We will include all SR articles irrespective of included primary study designs. The definition of SR is “a scientific investigation that focuses on a specific question and uses explicit, prespecified scientific methods to identify, select, assess, and summarize the findings of similar but separate studies.” (9). We will exclude Cochrane Reviews or the U.S. Preventive Services Task Force review because their backgrounds styles are prespecified by the respective organizations and different from usual original articles (10,11).

**Table 1.**
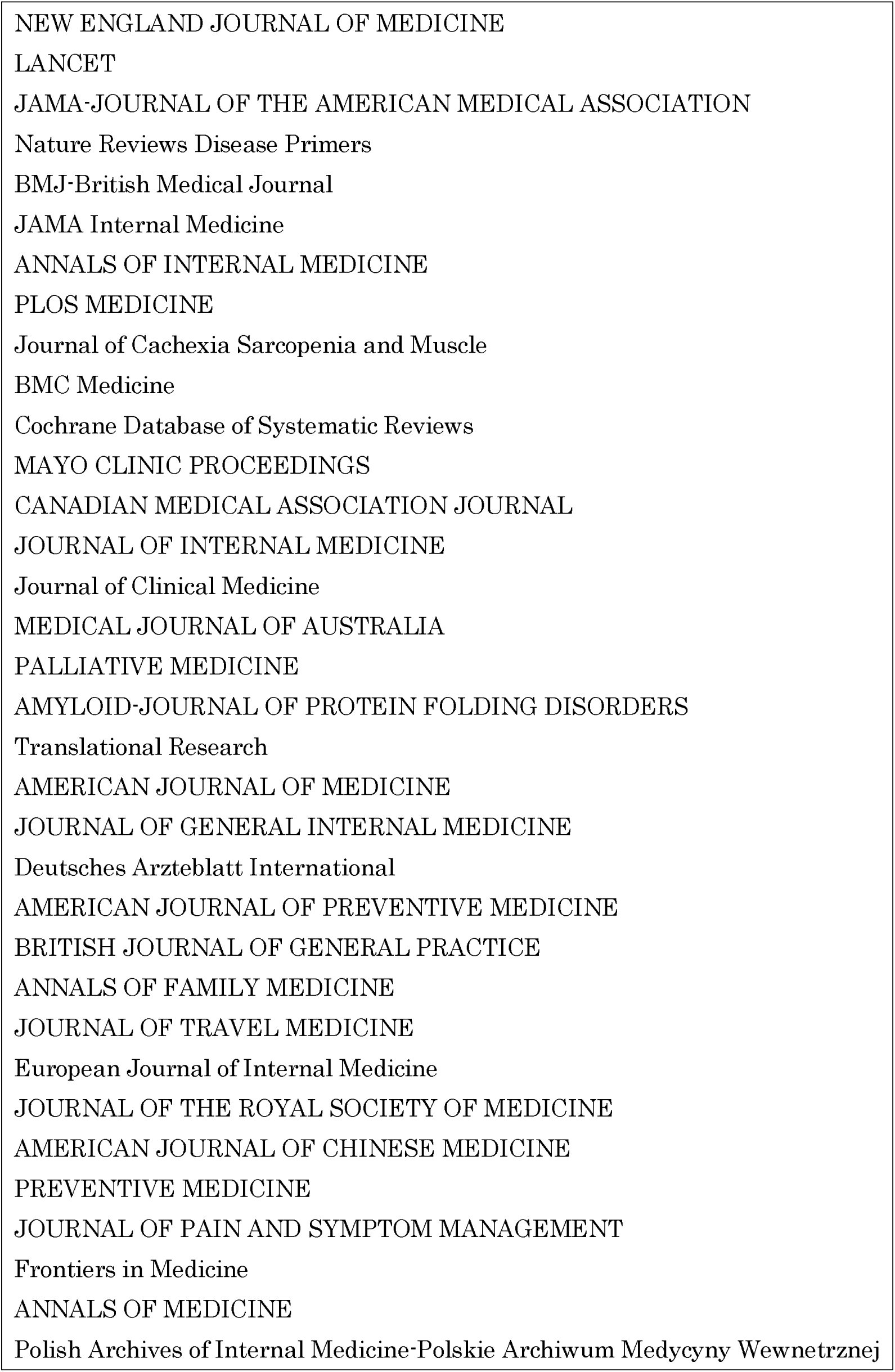
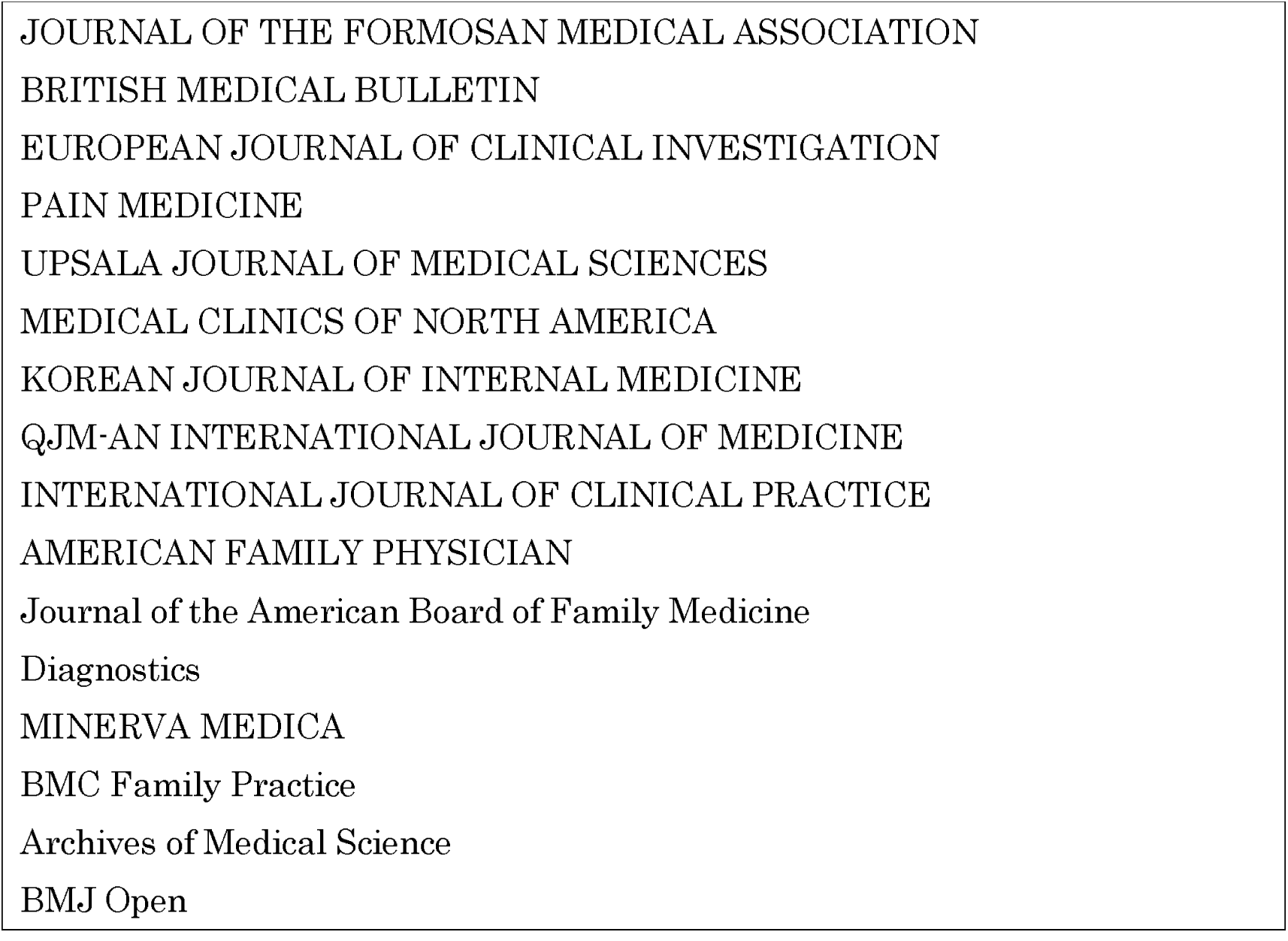
Top 50 journals in MEDICINE, GENERAL & INTERNAL category categorized in Journal Citation Reports 2018

### Information Sources

PubMed, Web of Science

### Search

We will search PubMed. The details of search formula are shown in Table 2A, and 2B.

**Table 2A.** Search formula of top 10 journals

|  | formula | number of references |
| --- | --- | --- |
| #1 | 2018[pdat] | 1329623 |
| #2 | "Systematic Review"[Publication Type] | 117216 |
| #3 | "N Engl J Med"[journal] OR "Lancet"[journal] OR "JAMA"[journal] OR "Nat Rev Dis Primers"[journal] OR "BMJ"[journal] OR "JAMA Internal Medicine"[journal] OR "Ann Intern Med"[journal] OR "PLoS Med"[journal] OR "J Cachexia Sarcopenia Muscle"[journal] OR "BMC Med"[journal] | 412907 |
| #4 | #1 AND #2 AND #3 | 131 |

**Table 2B.** Search formula of top 12 to 50 journals*

|  | formula | number of references |
| --- | --- | --- |
| #1 | 2018[pdat] | 1329623 |
| #2 | "Systematic Review"[Publication Type] | 117216 |
| #3 | "Mayo Clin Proc"[journal] OR "Can Med Assoc J"[journal] OR "J Intern Med"[journal] OR "J Clin Med"[journal] OR "Med J Aust"[journal] OR "Palliat Med"[journal] OR "Amyloid"[journal] OR "Transl Res"[journal] OR "Am J Med"[journal] OR "J Gen Intern Med"[journal] OR "Dtsch Arztebl Int"[Journal][journal] OR "Am J Prev Med"[journal] OR "Br J Gen Pract"[journal] OR "Ann Fam Med"[journal] OR "J Travel Med"[journal] OR "Eur J Intern Med"[journal] OR "J R Soc Med"[journal] OR "Am J Chin Med"[journal] OR "Prev Med"[journal] OR "J Pain Symptom Manage"[journal] OR "Front Med (Lausanne)"[journal] OR "Ann Med"[journal] OR "Pol Arch Intern Med"[journal] OR "J Formos Med Assoc"[journal] OR "Br Med Bull"[journal] OR "Eur J Clin Invest"[journal] OR "Pain Med"[journal] OR "Ups J Med Sci"[journal] OR "Med Clin North Am"[journal] OR "Korean J Intern Med"[journal] OR "QJM"[journal] OR "Int J Clin Pract"[journal] OR "Am Fam Physician"[journal] OR "J Am Board Fam Med"[journal] OR "Diagnostics (Basel)"[journal] OR "Minerva Med"[journal] OR "BMC Fam Pract"[journal] OR "Arch Med Sci"[journal] OR "BMJ Open"[journal] | 291774 |
| #4 | #1 AND #2 AND #3 | 429 |

\*excluded Cochrane Database of Systematic Reviews (11th ranking) search date 2019/12/01

### Study selection

One review author (YK) will confirm whether the articles are SR or not.

### Exposures

Four aspects that one can emphasize about one’s research question in the background section may be summarized as follows: “novelty”, “scope”, “quality”, and “update”. “Novelty” means a completely new research question.

“Scope” means that there are reports related to the research question, but authors expanded or limited the PICO.

“Quality” means that there are reports related to the research question, but there were methodological flaws.

“Update” means that there were same reports, but the search date was new.

We will conduct content analysis of the first 10 articles independently by four review authors (YK, ST, YT, or HY). We will develop a detailed guide from the initial review. We will resolve disagreements through the discussion, after that two of four review authors will conduct content analysis of the rest. We will resolve disagreements through the discussion. If necessary, another third reviewer will act as an arbiter. We will add other categories through the review if necessary. We will assess the agreement with kappa values.

### Primary outcome

The primary outcome will be whether the article was published in top 10 journal or not.

### Data items

Details are shown in Table 3. Considering that papers published in top journals will have many citations, we defined confoundings following previous studies which investigated the prognostic factors for increased citations (12–15). We will retrieve some data from Web of Science application programming interface using Python 3.7.4 software program (Python Software Foundation, De, USA).

**Table 3.**
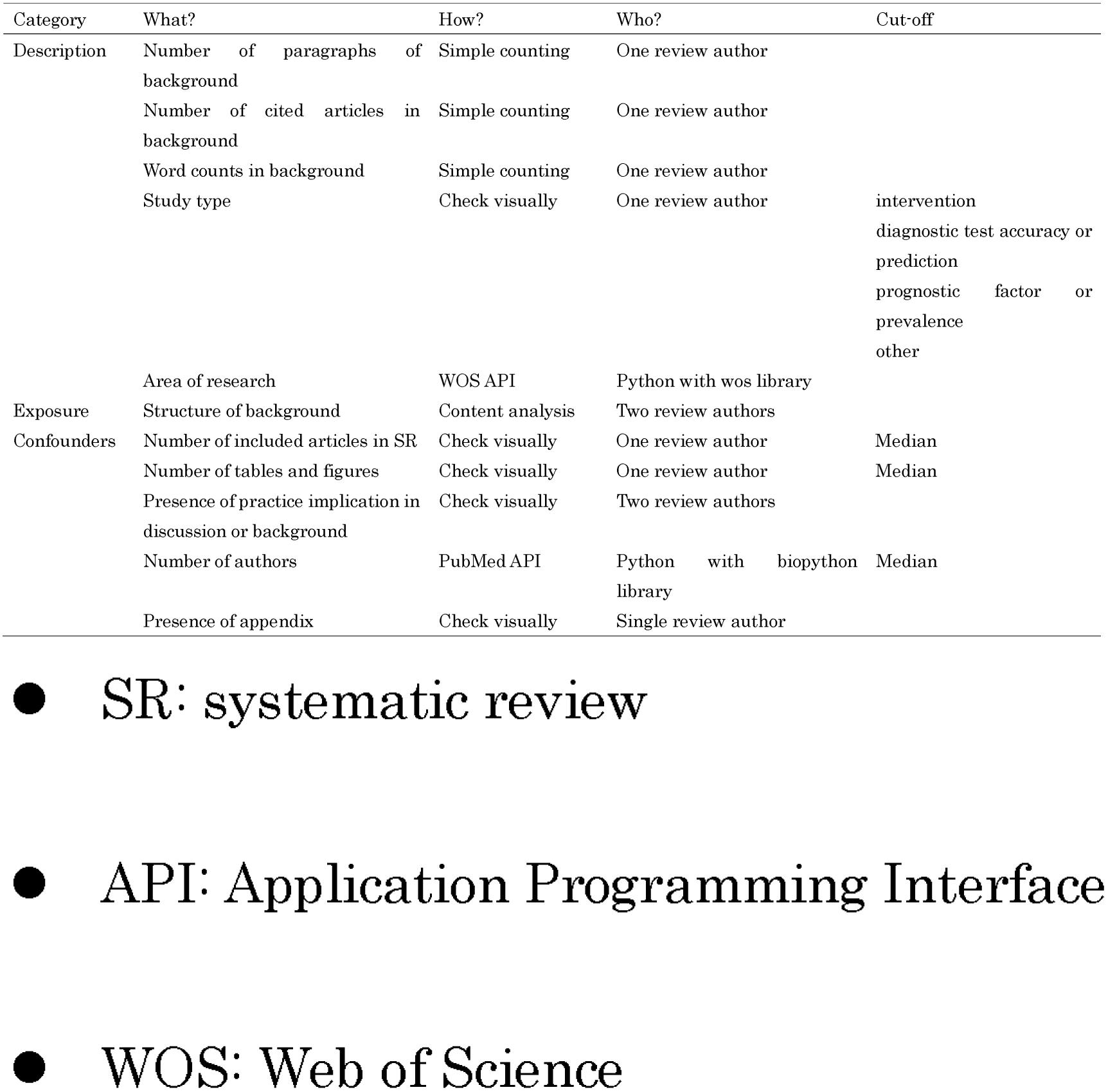
data items

### Statistical analysis

We will use descriptive statistics to summarize. We will use chi-squared test and logistic regression analysis. The primary analysis will be logistic regression predicting publication in high impact journals, with covariates as listed above. Two-tailed p values will be considered statistically significant if less than 0.05. We will use Stata ver. 16.0 (StataCorp LLC, College Station, Texas, United States of America). We determined the sample size as below: we will use 4 exposures and 5 confoundings. We need 90 events for the validity of the logistic model (16). We will randomly select 90 articles from the top 10 journals and 90 articles from the 11^th^ to 50^th^ journal as control.

## Discussion

This is the first study to investigate the influence of what to present and not present in the backgrounds section to be accepted in the highly cited journals among SR articles. There are several limitations. We will not assess the methodological quality of each articles due to the difficulty to score the quality in single measurement. For example, AMSTAR 2 (17), which is the most famous assessment tool, only accounts for intervention SRs. We will not take into account the clinical significance of the review, which is a confounding factor, but it is difficult to evaluate on one scale.

The results of this study will be a good help for systematic review authors not only when they write the background section, but also when they think about research questions.

- SR: systematic review
- API: Application Programming Interface
- WOS: Web of Science

## Data Availability

We will share the data related to this study, if requested.

